# Mitochondrial DNA copy number and incident heart failure: The Atherosclerosis Risk in Communities (ARIC) study

**DOI:** 10.1101/19012013

**Authors:** Yun Soo Hong, Ryan J. Longchamps, Di Zhao, Christina A. Castellani, Laura R. Loehr, Patricia P. Chang, Kunihiro Matsushita, Megan L. Grove, Eric Boerwinkle, Dan E. Arking, Eliseo Guallar

**Affiliations:** Departments of Epidemiology and Medicine, and Welch Center for Prevention, Epidemiology, and Clinical Research, Johns Hopkins University Bloomberg School of Public Health. Baltimore, Maryland, USA; McKusick-Nathans Department of Genetic Medicine, Johns Hopkins University School of Medicine, Baltimore, Maryland, USA; Department of Medicine, University of North Carolina School of Medicine, Chapel Hill, North Carolina, USA; Human Genetics Center, Department of Epidemiology, Human Genetics and Environmental Sciences, School of Public Health, The University of Texas Health Science Center at Houston, Houston, Texas, USA; Human Genome Sequencing Center, Baylor College of Medicine, Houston, Texas, USA

**Keywords:** cohort study, heart failure, mitochondrial DNA, mitochondrial DNA copy number, mitochondrial dysfunction

## Abstract

**Background:** The association between mitochondrial DNA-copy number (mtDNA-CN) and incident heart failure (HF) in the general population is unclear.

**Methods:** We examined the association between mtDNA-CN and the risk of incident HF among 10,802 participants free of HF at baseline from the Atherosclerosis Risk in Communities (ARIC) study, a large bi-racial population-based cohort. mtDNA-CN was estimated using probe intensities on the Affymetrix Genome-Wide Human single nucleotide polymorphisms Array 6.0. Incident HF events were identified through hospital discharge codes from 1987 until 2005 and through adjudication by the ARIC HF Classification Committee since 2005.

**Results:** During a median follow-up of 23.1 years, there were 2,227 incident HF events (incidence rate 10.3 per 1000 person-years). In fully adjusted models, the hazard ratios (95% confidence intervals) for HF comparing the 2^nd^ through 5^th^ quintiles of mtDNA-CN to the 1^st^ quintile were 0.91 (0.80–1.04), 0.82 (0.72–0.93), 0.81 (0.71–0.92), and 0.74 (0.65–0.85), respectively (*P* for trend < 0.001). In stratified analyses, the associations between mtDNA-CN and HF were similar across examined subgroups. The inverse association between mtDNA-CN and incident HF was stronger in HF with reduced ejection fraction (HFrEF) than in HF with preserved ejection fraction (HFpEF).

**Conclusions:** In this prospective cohort, mtDNA-CN was inversely associated with the risk of incident HF suggesting that reduced levels of mtDNA-CN, a biomarker of mitochondrial dysfunction, could reflect early susceptibility to HF.

## INTRODUCTION

Heart failure (HF) is a leading clinical and public health concern affecting 23 million people globally and 6.2 million adults in the United States alone.^1,2^ The lifetime risk of developing HF is 20% and the prevalence of HF in the US is expected to increase by nearly 50% by the year 2030.^3,4^ Despite recent improvements, the prognosis of HF is still poor, with 5-year mortality >40%.^5-8^ Thus, it is critical to identify novel pathways that can help design new preventive strategies and characterize subjects at high risk of developing HF.

Mitochondria generate nearly all energy used by the cell as adenosine triphosphate (ATP).^9^ Each mitochondrion has 2 to 10 copies of mitochondrial DNA (mtDNA), for a total of 10^3^ to 10^4^ copies of mtDNA per cell. mtDNA copy number (mtDNA-CN) changes with energy demands and with oxidative stress, and has been established as an indirect biomarker of mitochondrial dysfunction.^10,11^ Reduced mtDNA-CN measured in peripheral blood is associated with cardiovascular disease (CVD), all-cause mortality, hypertension, diabetes, chronic kidney disease, and sudden cardiac death.^12-18^

In a case-control study, hospitalized patients with HF had lower mtDNA-CN in peripheral blood compared to controls without HF, and when HF cases were followed up, those with lower mtDNA-CN had a higher risk of cardiovascular death and rehospitalization compared to those with higher mtDNA-CN.^19^ In small case-control studies, depletion of mtDNA in heart tissue samples was associated with HF.^20,21^ The association between mtDNA-CN and incident HF in the general population, however, is unknown. In the present study, we examined the association between baseline mtDNA-CN and the risk of incident HF in the Atherosclerosis Risk in Communities (ARIC) study, a large bi-racial population-based cohort.

## MATERIALS AND METHODS

### Study population

The ARIC study is a population-based prospective cohort of 15,792 individuals 45–65 years of age at the time of recruitment (1987–1989; Visit 1). ARIC participants were recruited from 4 US communities: Forsyth County, NC; Jackson, MS; suburban Minneapolis, MN; and Washington County, MD.^22^ Since the first study visit, there have been 6 subsequent in-person visits (visits 2–7, with visit 7 currently underway) and regular telephone interviews (annually and then semiannually since 2012). Our analysis was restricted to 11,453 White or Black participants who had DNA collected in one of the visits to generate mtDNA-CN measurements (**Supplementary Figure 1**). We then excluded Black participants recruited from Minnesota or Maryland (n = 1), participants without follow-up information (n = 1), and participants with prevalent HF at the time of DNA collection (n = 596). We further excluded participants missing information on body mass index (n = 17) and high-density lipoprotein (HDL) cholesterol (n = 14). The final sample included 10,802 participants (4,918 men and 5,904 women without HF at the time of DNA sampling). All centers obtained approval from their respective institutional review boards and all participants provided written informed consent.

### Measurements

ARIC participants underwent a comprehensive cardiovascular examination and interview by trained clinical staff members during each clinic visit. Age, sex, race/ethnicity, smoking status, alcohol intake, and medication history were self-reported. Smoking and alcohol intake were categorized as nxever, former, and current. Body mass index (BMI) was calculated from measured height and weight and categorized as underweight/normal (<25 kg/m^2^), overweight (≥25 to <30 kg/m^2^), or obese (≥30 kg/m^2^). Blood samples were collected for glucose, total cholesterol, and HDL-cholesterol measurements.

Hypertension was defined as systolic blood pressure ≥140 mmHg, diastolic blood pressure ≥90 mmHg, or current use of anti-hypertensive medication. Diabetes was defined as self-reported physician diagnosis of diabetes, fasting glucose ≥126 mg/dL, non-fasting glucose ≥200 mg/dL, or use of hypoglycemic medication. Prevalent coronary heart disease (CHD) was defined as the presence of a myocardial infarction based on self-report or electrocardiogram in visit 1, or the development of an adjudicated definite or probable myocardial infarction prior to the time of DNA collection for mtDNA-CN measurement.

### Measurement of mtDNA copy number

The methods for measuring mtDNA-CN have been described previously.^17,23^ Briefly, DNA was extracted using the Gentra Puregene Blood Kit (Qiagen N.V., Venlo, The Netherlands) from buffy coat of whole blood samples collected in visits 1–4. mtDNA-CN was calculated from probe intensities of mitochondrial single nucleotide polymorphisms (SNP) on the Affymetrix Genome-Wide Human SNP Array 6.0 using the Genvisis software package (www.genvisis.org), which uses the median mitochondrial probe intensity of 25 high-quality mitochondrial probes as initial raw measure of mtDNA-CN. Batch effects, DNA quality, and starting DNA quantity were corrected for by using surrogate variable analysis applied to probe intensities of 43,316 autosomal SNPs.^24^ The mtDNA-CN metric used in this analysis was obtained as the standardized residuals (mean 0 and standard deviation 1) in a linear regression in which initial raw estimates of mtDNA-CN were regressed against age, sex, enrollment center, surrogate variables used in the surrogate variable analysis, and white blood cell count. White blood cell count was not available in 14.9% of individuals and we imputed the mean for the missing values.^23^ DNA for mtDNA-CN for this analysis was obtained from ARIC visit 1 (1987– 1989) in 429 participants, visit 2 (1990–1992) in 8,655 participants, visit 3 (1993–1995) in 1,654 participants, and visit 4 (1996–1998) in 64 participants. For each participant, we used the visit in which DNA for mtDNA-CN assays was obtained as the baseline visit.

### Outcome definition and adjudication

Incident HF was defined as the first hospitalization for HF or death related to HF after the visit in which DNA was obtained for mtDNA-CN assays. Hospitalizations and deaths related to HF were identified as International Classification of Disease, 9^th^ Revision code 428, and International Classification of Diseases, 10^th^ Revision code I50 in discharge codes or in underlying cause of death, respectively. Since 2005, ARIC began adjudication of HF events by the ARIC HF Classification Committee.^25^ In addition, when available, adjudicated incident HF was further classified as HF with reduced ejection fraction (HFrEF, most recent left ventricular ejection fraction [LVEF] <50%), HF with preserved ejection fraction (HFpEF, LVEF ≥50%), or HF with unknown LVEF.

### Statistical analysis

Study participants were followed from the visit of DNA collection for mtDNA-CN assay until the development of HF, death, loss to follow-up, or December 31, 2017, whichever came first. We used a Cox proportional hazards model to estimate hazard ratios (HR) and 95% confidence intervals (CI) for the association between mtDNA-CN and incident HF. mtDNA-CN was first categorized into quintiles based on the overall sample distribution. In secondary analysis, mtDNA-CN was modeled as a continuous variable to estimate the HR for incident HF comparing the 90^th^ to the 10^th^ percentile of mtDNA-CN. In addition, mtDNA-CN was modeled as restricted quadratic splines with knots at the 5^th^, 50^th^, and 95^th^ percentiles to provide a smooth and flexible description of the dose-response relationship between mtDNA-CN and HF. We tested for the proportional hazards assumption using Schoenfeld residuals but the assumption was not met (*P* < 0.001). Therefore, to allow the effect of mtDNA-CN on incident HF to vary by time, we used a parametric survival models with spline variables created separately for baseline hazard and for time-dependent effects.^26^

To control for potential confounders, we used 4 models with progressive degrees of adjustment using covariates measured at the time of mtDNA-CN measurement: Model 1 was adjusted for age, sex, race, and enrollment center; Model 2 was further adjusted for BMI, smoking, and alcohol intake; Model 3 was further adjusted for total and HDL-cholesterol, cholesterol medication, hypertension, and diabetes; and Model 4 was further adjusted for prevalent CHD.

We performed stratified analyses by pre-specified subgroups defined by age (<60 or ≥60 years), sex, race (White or Black), smoking status (never, former, or current), alcohol intake (never, former, current), BMI (underweight/normal, overweight, or obese), and prevalent CHD status. We also performed several sensitivity analyses. First, we repeated the main analyses treating non-HF related deaths as a competing event using a proportional sub-distribution hazards model. Second, we used alternative definitions for incident HF: 1) discharge codes for HF for the entire follow-up period, regardless of adjudication; 2) HF events restricted to adjudicated cases that occurred since January 1, 2005, and 3) discharge codes for HF restricted to cases that occurred since January 1, 2005. For the two analyses restricted to events since 2005, we used late entries to address the issue of immortal person time prior to 2005. Third, also using HF events adjudicated after 2005 and late entries, we estimated the HRs for HFrEF and HFpEF separately using a proportional sub-distribution hazards model with non-HF related deaths and unknown type of HF as competing events. All statistical analyses were performed using Stata version 15.0 (StataCorp LP, College Station, TX, USA).

## RESULTS

The mean age (standard deviation) of study participants at baseline was 57.3 (5.9) years (**Table 1**). Participants with lower mtDNA-CN levels were more likely to be current smokers, to have a higher prevalence of hypertension, diabetes, and CHD, and to have lower HDL-cholesterol levels than those with higher mtDNA-CN levels.

**Table 1.** Baseline characteristics of study participants by quintile of mitochondrial DNA copy number.

|  | **Overall**<br>**(n = 10,802)** | **Quintile 1**<br>**(n = 2,161)** | **Quintile 2**<br>**(n = 2,160)** | **Quintile 3**<br>**(n = 2,161)** | **Quintile 4**<br>**(n = 2,160)** | **Quintile 5**<br>**(n = 2,160)** | **P-values<sup>†</sup>** |
| --- | --- | --- | --- | --- | --- | --- | --- |
| **mtDNA-CN range** | -5.16, 4.84 | -5.16, -0.72 | -0.72, -0.16 | -0.16, 0.29 | 0.29, 0.78 | 0.78, 4.84 |  |
| **Age (year)** | 57.3 (5.9) | 57.3 (6.0) | 57.3 (6.1) | 57.3 (5.9) | 57.2 (5.9) | 57.3 (5.8) | 0.78 |
| **BMI (kg/m<sup>2</sup>)** | 27.8 (5.3) | 27.8 (5.4) | 27.6 (5.1) | 28.1 (5.4) | 27.7 (5.3) | 27.9 (5.3) | 0.78 |
| **Male (%)** | 45.4 | 45.2 | 44.9 | 46.3 | 44.5 | 46.2 | 0.71 |
| **Race (%)** |  |  |  |  |  |  | 0.61 |
| White | 79.2 | 80.3 | 79.1 | 78.9 | 78.3 | 79.3 |  |
| Black | 20.8 | 19.7 | 20.9 | 21.2 | 21.7 | 20.7 |  |
| **Smoking (%)** |  |  |  |  |  |  | 0.02 |
| Never | 38.6 | 36.6 | 40.2 | 39.0 | 38.8 | 38.5 |  |
| Former | 38.5 | 37.7 | 36.8 | 38.8 | 39.7 | 40.0 |  |
| Current | 22.7 | 25.6 | 23.1 | 22.2 | 21.5 | 21.5 |  |
| **Current drinker (%)** | 58.0 | 55.3 | 59.0 | 57.5 | 61.1 | 57.9 | 0.003 |
| **TC (mg/dl)** | 209.7 (39.4) | 209.5 (41.7) | 208.8 (39.3) | 209.3 (37.3) | 209.5 (38.0) | 211.6 (40.7) | 0.07 |
| **HDL-C (mg/dl)** | 49.8 (17.1) | 48.4 (16.8) | 49.5 (17.1) | 49.8 (16.7) | 51.2 (17.7) | 49.9 (17.2) | < 0.001 |
| **TG (mg/dl)\*** | 115 (83–162) | 117 (84–164.5) | 116 (83–162) | 115 (82–165) | 109 (81–158) | 118 (85–162) | 0.003 |
| **SBP (mmHg)** | 122.0 (19.1) | 122.7 (19.6) | 122.0 (19.9) | 122.0 (18.4) | 121.6 (18.9) | 121.6 (18.6) | 0.05 |
| **Prevalent CHD (%)** | 4.8 | 6.5 | 5.0 | 4.8 | 3.9 | 4.0 | 0.001 |
| **Hypertension (%)** | 34.6 | 37.0 | 35.1 | 33.6 | 33.2 | 34.5 | 0.08 |
| **Diabetes (%)** | 14.0 | 17.3 | 14.2 | 13.5 | 12.1 | 12.9 | < 0.001 |

Abbreviations: BMI, body mass index; CHD, coronary heart disease; HDL-C, high-density lipoprotein-cholesterol; mtDNA-CN, mitochondrial DNA copy number; SBP, systolic blood pressure; TC, total cholesterol; and TG, triglyceride.
\* Median (interquartile range).
<sup>†</sup> *P* for trend for continuous variables; *P* values from $\chi^2$ tests for categorical variables.

During a median follow-up of 23.1 years, we identified 2,227 new cases of HF (incidence rate of 10.3 per 1000 person-years). In fully adjusted models, the HRs (95% CI) for HF comparing the 2^nd^ through 5^th^ quintiles of mtDNA-CN to the 1^st^ quintile were 0.91 (0.80–1.04), 0.82 (0.72–0.93), 0.81 (0.71–0.92), and 0.74 (0.65–0.85), respectively (*P* for trend < 0.001; **Table 2**). The fully adjusted HR for HF comparing the 90^th^ to the 10^th^ percentile of mtDNA-CN was 0.76 (0.69–0.84). In spline regression analysis, mtDNA-CN was inversely associated with the risk of incident HF with an approximately linear dose-response relationship (P-value for non-linear spline terms 0.74; **Figure 1**). The results were similar when we used hospital discharge codes for HF, adjudicated HF cases since 2005, or hospital discharge codes for HF since 2005, or when non-HF related deaths were treated as a competing event (**Supplementary Tables 1–4**).

**Table 2.** Hazard ratios for incident heart failure by levels of mitochondrial DNA copy number.

| mtDNA-CN | N events / N total | Incidence rate per 1,000 | Model 1 | Model 2 | Model 3 | Model 4 |
| --- | --- | --- | --- | --- | --- | --- |
| **Overall** | 2,227 / 10,802 | 10.3 |  |  |  |  |
| **Quintile 1** | 526 / 2,161 | 12.7 | 1 (reference) | 1 (reference) | 1 (reference) | 1 (reference) |
| **Quintile 2** | 455 / 2,160 | 10.6 | 0.82 (0.73–0.93) | 0.89 (0.78–1.01) | 0.90 (0.80–1.03) | 0.91 (0.80–1.04) |
| **Quintile 3** | 427 / 2,161 | 9.9 | 0.77 (0.68–0.87) | 0.79 (0.69–0.89) | 0.81 (0.71–0.93) | 0.82 (0.72–0.93) |
| **Quintile 4** | 416 / 2,160 | 9.3 | 0.71 (0.63–0.81) | 0.77 (0.68–0.88) | 0.80 (0.70–0.91) | 0.81 (0.71–0.92) |
| **Quintile 5** | 403 / 2,160 | 9.2 | 0.70 (0.62–0.80) | 0.74 (0.65–0.84) | 0.73 (0.64–0.84) | 0.74 (0.65–0.85) |
| **P for trend** |  |  | < 0.001 | < 0.001 | < 0.001 | < 0.001 |
| **90<sup>th</sup> vs. 10<sup>th</sup> percentile** |  |  | 0.71 (0.65–0.78) | 0.73 (0.66–0.81) | 0.75 (0.68–0.82) | 0.76 (0.69–0.84) |

Abbreviations: mtDNA-CN, mitochondrial DNA copy number.
Model 1: Adjusted for age, sex, race, and enrollment center; Model 2: Model 1 + body mass index, smoking, and alcohol intake; Model 3: Model 2 + total cholesterol, HDL-cholesterol, cholesterol medication, hypertension, and diabetes; and Model 4: Model 3 + prevalent coronary heart disease.

**Figure 1.**
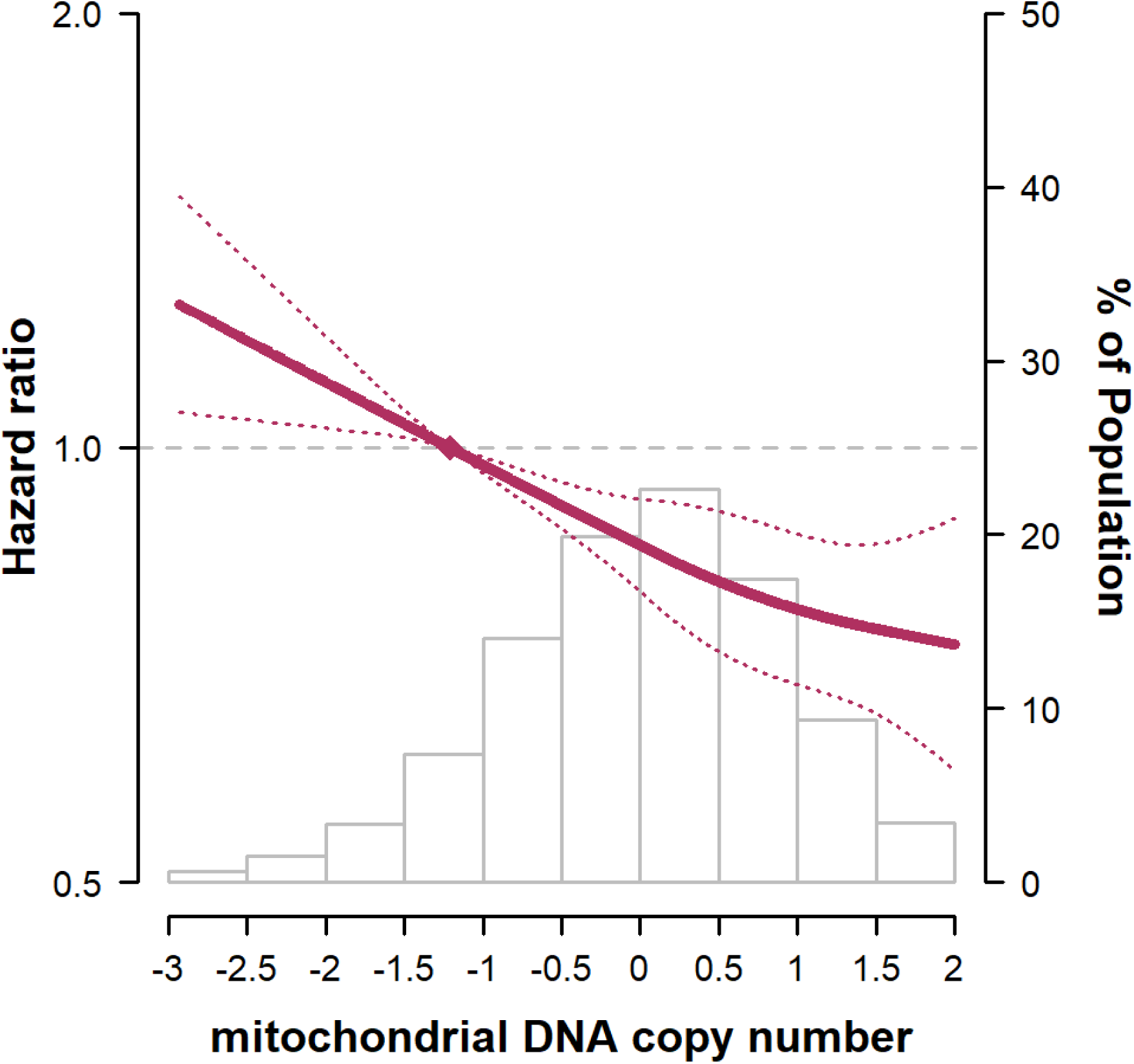
Hazard ratios for incident heart failure by levels of mitochondrial DNA copy number. The curves represent adjusted hazard ratios (solid line) and their 95% confidence intervals (dotted lines) based on restricted quadratic splines of mtDNA copy number with knots at 5^th^, 50^th^, and 95^th^ percentiles of its distribution. The reference value (diamond dot) was set at the 10^th^ percentile of the distribution. Results were obtained from a Cox model adjusted for age, sex, race/ethnicity, body mass index, smoking, alcohol intake, total and HDL cholesterol, cholesterol medication, hypertension, diabetes, and prevalent coronary heart disease. Histograms represent the frequency distribution of mtDNA copy number at baseline.

In stratified analyses, the associations between mtDNA-CN and HF were similar across subgroups defined by age, sex, race, smoking status, and history of CHD (**Figure 2**). The associations were weaker in participants who currently drink alcohol compared to those who did not drink (*P* for interaction = 0.01), and in participants who were overweight compared to those who were underweight or normal weight (*P* for interaction = 0.05).

**Figure 2.**
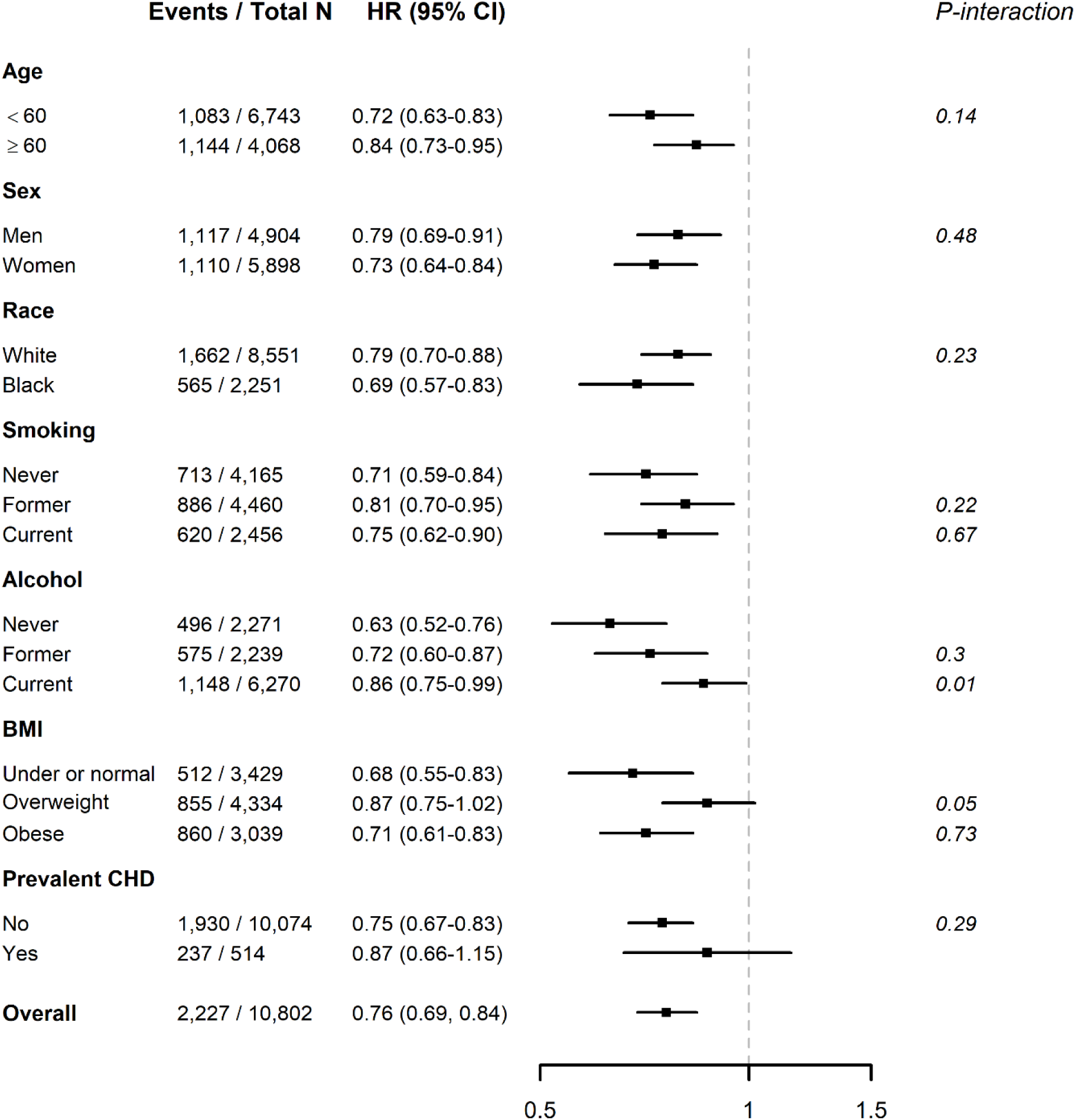
Hazard ratios for incident heart failure comparing the 90^th^ to the 10^th^ percentile of mitochondrial DNA copy number in selected subgroups. The figure includes hazard ratios for comparing the 90^th^ to the 10^th^ percentile (reference) of mtDNA copy number. Pre-specified subgroups were age (<60 or ≥60 years), sex, race (White or Black), smoking status (never, former, or current), alcohol intake (never, former, current), BMI (underweight/normal, overweight, or obese), and prevalent CHD. Models were adjusted for age, sex, race/ethnicity, body mass index, smoking, alcohol intake, total and HDL cholesterol, cholesterol medication, hypertension, diabetes, and prevalent coronary heart disease.

When we tested the proportional hazards assumption, the association between mtDNA-CN and HF was progressively attenuated from the time of mtDNA-CN measurement (**Table 3 and Figure 3**). The fully adjusted HRs for HF comparing the 90^th^ to the 10^th^ percentile of mtDNA-CN at 10, 20, and 30 years since mtDNA-CN measurement were 0.65 (0.54–0.79), 0.89 (0.76–1.04), and 0.99 (0.72–1.37), respectively.

**Table 3.** Hazard ratios for incident heart failure at 10, 20, and 30 years since mtDNA copy number measurement.

|  | **Year 10** | **Year 20** | **Year 30** |
| --- | --- | --- | --- |
| **Quintile 1** | 1 (reference) | 1 (reference) | 1 (reference) |
| **Quintile 2** | 0.90 (0.78–1.03) | 0.94 (0.83–1.08) | 0.97 (0.82–1.14) |
| **Quintile 3** | 0.78 (0.66–0.93) | 0.87 (0.75–1.02) | 0.91 (0.72–1.16) |
| **Quintile 4** | 0.76 (0.61–0.93) | 0.89 (0.74–1.06) | 0.95 (0.69–1.32) |
| **Quintile 5** | 0.68 (0.53–0.88) | 0.84 (0.69–1.02) | 0.92 (0.61–1.39) |
| **90th vs. 10th percentile** | 0.65 (0.54–0.79) | 0.89 (0.76–1.04) | 0.99 (0.72–1.37) |

Time-dependent model adjusted for age, sex, race, enrollment center, body mass index, smoking, alcohol intake, total cholesterol, HDL-cholesterol, cholesterol medication, hypertension, diabetes, and prevalent coronary heart disease.

**Figure 3.**
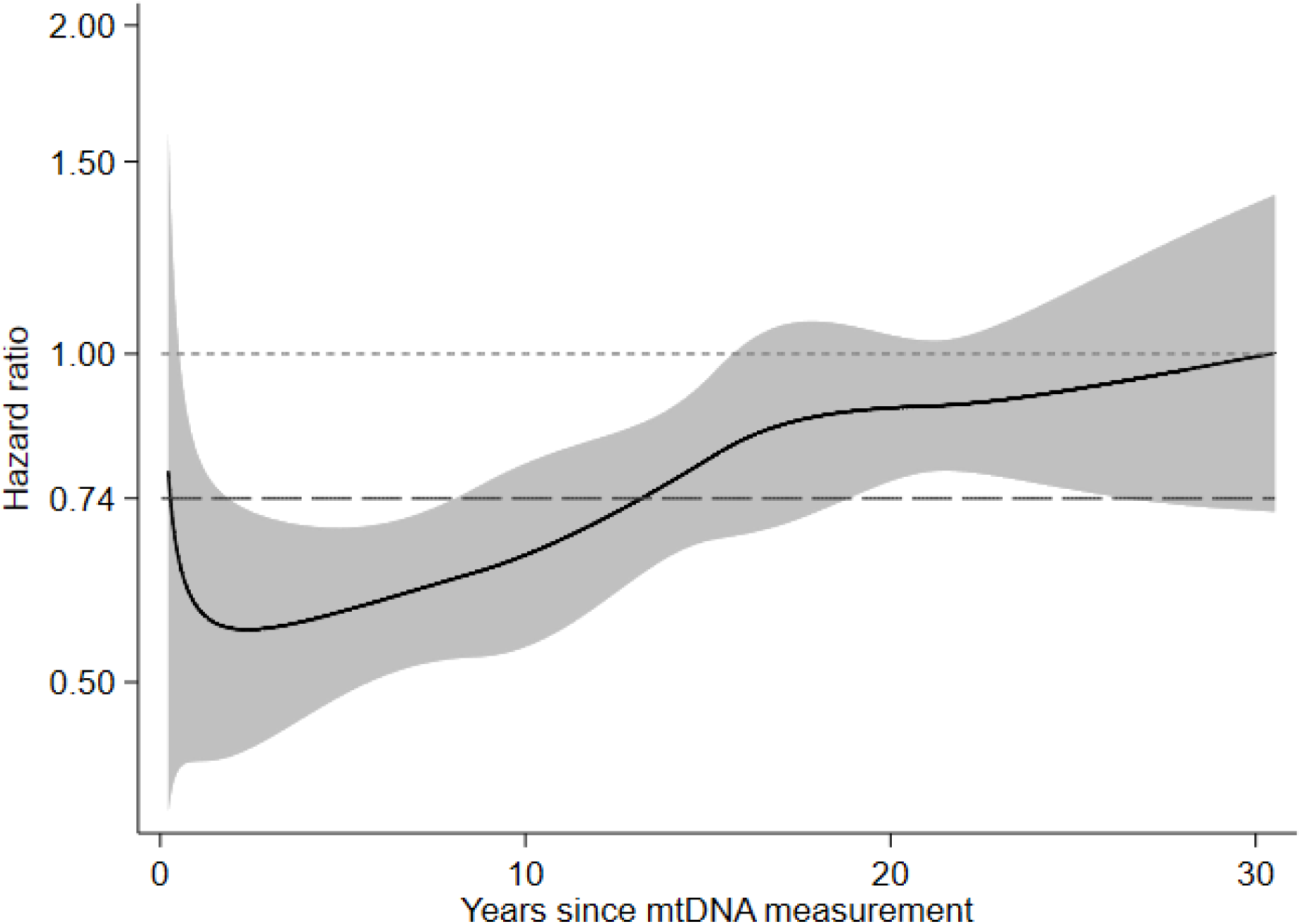
Time-dependent hazard ratios for incident heart failure comparing the 90^th^ to the 10^th^ percentile of mitochondrial DNA copy number. The curve represents time-dependent adjusted hazard ratios (solid line) and the gray band represents its corresponding 95% confidence interval comparing the 90^th^ to the 10^th^ percentile of mtDNA copy number. The dashed line represents the time-fixed adjusted hazard ratio comparing the 90^th^ to the 10^th^ percentile of mtDNA copy number. Models were adjusted for age, sex, race/ethnicity, body mass index, smoking, alcohol intake, total and HDL cholesterol, cholesterol medication, hypertension, diabetes, and prevalent coronary heart disease.

Finally, when we separated HF events into HFpEF and HFrEF using data since 2005, the inverse association between mtDNA-CN and incident HF was stronger in HFrEF than in HFpEF (**Table 4)**, although the trend was not statistically significant in either type of HF (*P* for trend 0.73 and 0.12 in HFpEF and HFrEF, respectively).

**Table 4.** Hazard ratios for incident heart failure with preserved ejection fraction (HFpEF) and heart failure with reduced ejection fraction (HFrEF) by quintiles of mitochondrial DNA copy number.

| mtDNA-CN | N events / N total | Incidence rate per 1,000 | Model 1 | Model 2 | Model 3 | Model 4 |
| --- | --- | --- | --- | --- | --- | --- |
| **HFpEF** |  |  |  |  |  |  |
| **Overall** | 564 / 8,892 | 6.2 |  |  |  |  |
| **Quintile 1** | 103 / 1,701 | 6.0 | 1 (reference) | 1 (reference) | 1 (reference) | 1 (reference) |
| **Quintile 2** | 111 / 1,776 | 6.1 | 1.05 (0.80–1.37) | 1.09 (0.83–1.43) | 1.10 (0.94–1.45) | 1.12 (0.85–1.47) |
| **Quintile 3** | 120 / 1,762 | 6.6 | 1.16 (0.89–1.51) | 1.16 (0.89–1.51) | 1.17 (0.90–1.53) | 1.17 (0.90–1.53) |
| **Quintile 4** | 122 / 1,846 | 6.2 | 1.12 (0.86–1.45) | 1.15 (0.88–1.49) | 1.17 (0.89–1.52) | 1.13 (0.86–1.48) |
| **Quintile 5** | 108 / 1,807 | 5.8 | 1.01 (0.77–1.32) | 1.02 (0.78–1.34) | 1.02 (0.77–1.34) | 0.97 (0.74–1.29) |
| **P for trend** |  |  | 0.80 | 0.76 | 0.77 | 0.89 |
| **HFrEF** |  |  |  |  |  |  |
| **Overall** | 504 / 8,892 | 5.5 |  |  |  |  |
| **Quintile 1** | 114 / 1,701 | 6.7 | 1 (reference) | 1 (reference) | 1 (reference) | 1 (reference) |
| **Quintile 2** | 103 / 1,776 | 5.7 | 0.87 (0.66–1.13) | 0.88 (0.68–1.16) | 0.88 (0.67–1.15) | 0.88 (0.67–1.16) |
| **Quintile 3** | 94 / 1,762 | 5.1 | 0.80 (0.64–1.05) | 0.81 (0.62–1.07) | 0.81 (0.62–1.07) | 0.79 (0.59–1.04) |
| **Quintile 4** | 100 / 1,846 | 5.1 | 0.82 (0.63–1.07) | 0.85 (0.64–1.11) | 0.85 (0.65–1.12) | 0.87 (0.66–1.15) |
| **Quintile 5** | 93 / 1,807 | 5.0 | 0.76 (0.58–1.00) | 0.77 (0.58–1.02) | 0.77 (0.58–1.02) | 0.80 (0.60–1.05) |
| **P for trend** |  |  | 0.06 | 0.07 | 0.08 | 0.14 |

Abbreviations: HFpEF, heart failure with preserved ejection fraction; HFrEF, heart failure with reduced ejection fraction; and mtDNA-CN, mitochondrial DNA copy number.
Model 1: Adjusted for age, sex, and race/enrollment center; Model 2: Model 1 + body mass index, smoking, alcohol intake, and physical activity; Model 3: Model 2 + total cholesterol, HDL-cholesterol, cholesterol medication, hypertension, diabetes; Model 4: Model 3 + prevalent coronary heart disease.

## DISCUSSION

In this large community-based prospective cohort, mtDNA-CN was inversely associated with the risk of incident HF. The association was approximately linear and consistent across population subgroups. The association, however, was strongest early after measurement of mtDNA-CN and was progressively attenuated over 30 years of follow-up. These findings suggest a potential role of mtDNA-CN as an early indicator for HF, particularly for events in the near future.

Decreased mtDNA-CN is associated with CVD events, including sudden cardiac death, all-cause mortality, hypertension, diabetes, and chronic kidney disease.^12-18^ However, the association between mtDNA-CN and incident HF is largely unknown. In animal studies, mtDNA damage and depletion were associated with the development of dilated cardiomyopathy and impaired left ventricular remodeling after ischemic injury.^27-30^ In small case-control studies, patients with HF had decreased mtDNA-CN levels in heart tissue samples compared to controls without HF.^21,31^ In another case-control study, hospitalized patients with HF had lower mtDNA-CN in peripheral blood compared to those without HF.^19^ When patients with HF were followed up for a median of 17 months, those with peripheral blood mtDNA-CN levels below the median were more likely to experience cardiovascular death and rehospitalization compared to those with mtDNA-CN levels above the median.^19^

The mechanisms underlying the association between mtDNA-CN and HF are unclear. Established risk factors for HF include hypertension, diabetes, metabolic syndrome, ischemic heart disease, and non-ischemic cardiomyopathies,^32,33^ and oxidative stress is assumed to play a major role in the development and progression of HF.^34-38^ Endogenous mtDNA damage is mainly caused by reactive oxygen species (ROS) as a by-product of oxidative phosphorylation. High levels of ROS trigger further increases in ROS generation, a process known as mitochondrial ROS-induced ROS release.^39^ Exogeneous agents, such as cigarette smoke, industrial by-products, ultraviolet and ionizing radiation, environmental toxins and chemicals, and chemotherapeutic drugs may also damage mtDNA.^40^ mtDNA is particularly susceptible to damage and mutations due to its close proximity to mitochondrial ROS production sites, lack of protective histones, and limited repair activity.^34^ Moreover, damage caused by ROS is more extensive and persists longer in mtDNA than in nuclear DNA.^41^

Mitochondrial ROS generation and mtDNA damage in cardiomyocytes lead to impaired electron chain transport and ATP synthesis, modifications to proteins involved in excitation-contraction coupling, and activation of hypertrophy signaling kinases and transcription factors, apoptotic pathways, inflammatory mediators, and matrix metalloproteinases.^34,42,43^ This process leads to a reduction in the number of mitochondria, hypertrophy, apoptosis, and necrosis of cardiomyocytes, impairment of contractile function, and fibrosis, which, in combination, lead to the initiation and progression of cardiac remodeling and ultimately to HF. Systemic oxidative stress is also closely linked to the development of vascular diseases, such as hypertension, which are major risk factors for HF.^34,38,44,45^ Finally, treatment with chemotherapeutic agents that inhibit DNA replication, such as nucleoside reverse transcriptase inhibitors or anthracycline, is also associated with both mtDNA depletion and cardiomyopathy.^46-48^

In our study, mtDNA-CN measurements were derived from the buffy coat of peripheral blood and were not a direct measurement of mtDNA-CN in cardiomyocytes. Leukocyte mtDNA-CN was correlated with mtDNA-CN in cardiomyocytes in one study.^19^ In another study of non-ischemic cardiomyopathy patients, there was a moderate correlation between whole blood mtDNA-CN and myocardial mtDNA-CN.^49^ Additional research is needed to understand the association between mtDNA-CN in peripheral blood and in target tissues, and to elucidate the mechanisms linking mtDNA-CN in peripheral blood with incident HF.

Other limitations also need to be considered in the interpretation of our findings. First, mtDNA-CN was measured only once and we could not evaluate changes in mtDNA-CN after the baseline visit. In fact, the association between mtDNA-CN and incident HF decreased progressively over follow-up, although mtDNA-CN was inversely associated with HF for at least two decades after measurement. Second, systematic adjudication of HF events only occurred after 2005. However, the sensitivity and positive predictive value of ICD-9 codes for HF compared to adjudicated acute decompensated HF and chronic HF in ARIC were relatively high at 0.95 and 0.77, respectively, and the results from our sensitivity analyses were consistent for analyses based on discharge codes and for those based on adjudicated events.^25^ Third, our analysis of the specific association of mtDNA-CN levels and subtypes of HF was restricted to events occurring after 2005, and we had limited power to identify differences between HFpEF and HFrEF. Finally, we were not able to evaluate the association between mtDNA-CN level and the severity of HF symptoms as such information was not available for all participants.

The major strengths of this study include the prospective design with a long duration of follow-up to capture HF events, the large sample size, the high quality of field and laboratory methods of the ARIC study, and the ability to account for multiple potential confounders for the association between mtDNA-CN and incident HF. In addition, we used state-of-the art tools to measure mtDNA-CN.^23^

## CONCLUSIONS

In this large community-based prospective cohort, mtDNA-CN was inversely associated with the risk of incident HF suggesting that reduced levels of mtDNA-CN, a biomarker of mitochondrial dysfunction, could reflect early susceptibility to HF. Further studies are needed to better understand the underlying mechanisms and to characterize the association of mtDNA-CN with different types of HF and their severity.

## Data Availability

The detailed policies for requesting and accessing the ARIC data can be found in the following website: https://sites.cscc.unc.edu/aric/distribution-agreements. It is also possible to obtain most ARIC data from BioLINCC, a repository maintained by the National Heart, Lung, and Blood Institute. The BioLINCC website (https://biolincc.nhlbi.nih.gov/) includes detailed information about the available data and the process to obtain such data.

## ACKNOWLEDGEMENTS

The authors thank the staff and participants of the ARIC study for their important contributions.

## REFERENCES

1. Bui AL, Horwich TB, Fonarow GC. Epidemiology and risk profile of heart failure. Nature reviews Cardiology 2011;8:30–41.

2. Benjamin EJ, Muntner P, Alonso A, et al. Heart Disease and Stroke Statistics—2019 Update: A Report From the American Heart Association. Circulation 2019;139:e56–e528.

3. Metra M, Teerlink JR. Heart failure. Lancet (London, England) 2017;390:1981–95.

4. Lloyd-Jones DM, Larson MG, Leip EP, et al. Lifetime risk for developing congestive heart failure: the Framingham Heart Study. Circulation 2002;106:3068–72.

5. Crespo-Leiro MG, Anker SD, Maggioni AP, et al. European Society of Cardiology Heart Failure Long-Term Registry (ESC-HF-LT): 1-year follow-up outcomes and differences across regions. European journal of heart failure 2016;18:613–25.

6. Chang PP, Chambless LE, Shahar E, et al. Incidence and survival of hospitalized acute decompensated heart failure in four US communities (from the Atherosclerosis Risk in Communities Study). The American journal of cardiology 2014;113:504–10.

7. Loehr LR, Rosamond WD, Chang PP, Folsom AR, Chambless LE. Heart failure incidence and survival (from the Atherosclerosis Risk in Communities study). The American journal of cardiology 2008;101:1016–22.

8. Gerber Y, Weston SA, Redfield MM, et al. A contemporary appraisal of the heart failure epidemic in Olmsted County, Minnesota, 2000 to 2010. JAMA internal medicine 2015;175:996–1004.

9. Friedman JR, Nunnari J. Mitochondrial form and function. Nature 2014;505:335–43.

10. Clay Montier LL, Deng JJ, Bai Y. Number matters: control of mammalian mitochondrial DNA copy number. Journal of genetics and genomics = Yi chuan xue bao 2009;36:125–31.

11. Malik AN, Czajka A. Is mitochondrial DNA content a potential biomarker of mitochondrial dysfunction? Mitochondrion 2013;13:481–92.

12. Zhang Y, Guallar E, Ashar FN, et al. Association between mitochondrial DNA copy number and sudden cardiac death: findings from the Atherosclerosis Risk in Communities study (ARIC). European heart journal 2017;38:3443–8.

13. Ashar FN, Zhang Y, Longchamps RJ, et al. Association of Mitochondrial DNA Copy Number With Cardiovascular Disease. JAMA cardiology 2017;2:1247–55.

14. Ashar FN, Moes A, Moore AZ, et al. Association of mitochondrial DNA levels with frailty and all-cause mortality. Journal of molecular medicine (Berlin, Germany) 2015;93:177–86.

15. Tang X, Luo YX, Chen HZ, Liu DP. Mitochondria, endothelial cell function, and vascular diseases. Frontiers in physiology 2014;5:175.

16. Lee HK, Song JH, Shin CS, et al. Decreased mitochondrial DNA content in peripheral blood precedes the development of non-insulin-dependent diabetes mellitus. Diabetes research and clinical practice 1998;42:161–7.

17. Tin A, Grams ME, Ashar FN, et al. Association between Mitochondrial DNA Copy Number in Peripheral Blood and Incident CKD in the Atherosclerosis Risk in Communities Study. Journal of the American Society of Nephrology : JASN 2016;27:2467–73.

18. Yue P, Jing S, Liu L, et al. Association between mitochondrial DNA copy number and cardiovascular disease: Current evidence based on a systematic review and meta-analysis. PloS one 2018;13:e0206003.

19. Huang J, Tan L, Shen R, Zhang L, Zuo H, Wang DW. Decreased Peripheral Mitochondrial DNA Copy Number is Associated with the Risk of Heart Failure and Long-term Outcomes. Medicine 2016;95:e3323.

20. Karamanlidis G, Nascimben L, Couper GS, Shekar PS, del Monte F, Tian R. Defective DNA replication impairs mitochondrial biogenesis in human failing hearts. Circulation research 2010;106:1541–8.

21. Karamanlidis G, Bautista-Hernandez V, Fynn-Thompson F, Nido Pd, Tian R. Impaired Mitochondrial Biogenesis Precedes Heart Failure in Right Ventricular Hypertrophy in Congenital Heart Disease. 2011;4:707–13.

22. The Atherosclerosis Risk in Communities (ARIC) Study: design and objectives. The ARIC investigators. American journal of epidemiology 1989;129:687–702.

23. Longchamps R, Castellani C, Newcomb C, et al. Evaluation of mitochondrial DNA copy number estimation techniques. bioRxiv 2019:610238.

24. Leek JT, Johnson WE, Parker HS, Jaffe AE, Storey JD. The sva package for removing batch effects and other unwanted variation in high-throughput experiments. Bioinformatics (Oxford, England) 2012;28:882–3.

25. Rosamond WD, Chang PP, Baggett C, et al. Classification of heart failure in the atherosclerosis risk in communities (ARIC) study: a comparison of diagnostic criteria. Circulation Heart failure 2012;5:152–9.

26. Royston P, Lambert PC. Flexible parametric survival analysis using Stata: beyond the Cox model: Stata College Station, Texas; 2011.

27. Ide T, Tsutsui H, Hayashidani S, et al. Mitochondrial DNA damage and dysfunction associated with oxidative stress in failing hearts after myocardial infarction. Circulation research 2001;88:529–35.

28. Ikeuchi M, Matsusaka H, Kang D, et al. Overexpression of mitochondrial transcription factor a ameliorates mitochondrial deficiencies and cardiac failure after myocardial infarction. Circulation 2005;112:683–90.

29. Wang J, Wilhelmsson H, Graff C, et al. Dilated cardiomyopathy and atrioventricular conduction blocks induced by heart-specific inactivation of mitochondrial DNA gene expression. Nature genetics 1999;21:133–7.

30. Kuznetsova T, Knez J. Peripheral Blood Mitochondrial DNA and Myocardial Function. In: Santulli G, ed. Mitochondrial Dynamics in Cardiovascular Medicine. Cham: Springer International Publishing; 2017:347-58.

31. Ahuja P, Wanagat J, Wang Z, et al. Divergent mitochondrial biogenesis responses in human cardiomyopathy. Circulation 2013;127:1957–67.

32. Yancy CW, Jessup M, Bozkurt B, et al. 2013 ACCF/AHA guideline for the management of heart failure: a report of the American College of Cardiology Foundation/American Heart Association Task Force on Practice Guidelines. Journal of the American College of Cardiology 2013;62:e147–239.

33. Ponikowski P, Voors AA, Anker SD, et al. 2016 ESC Guidelines for the diagnosis and treatment of acute and chronic heart failure: The Task Force for the diagnosis and treatment of acute and chronic heart failure of the European Society of Cardiology (ESC)Developed with the special contribution of the Heart Failure Association (HFA) of the ESC. European heart journal 2016;37:2129–200.

34. Tsutsui H, Kinugawa S, Matsushima S. Oxidative stress and heart failure. American journal of physiology Heart and circulatory physiology 2011;301:H2181–90.

35. Tsutsui H, Kinugawa S, Matsushima S. Oxidative Stress and Mitochondrial DNA Damage in Heart Failure. Circulation Journal 2008;72:A31–A7.

36. Takimoto E, Kass DA. Role of oxidative stress in cardiac hypertrophy and remodeling. Hypertension (Dallas, Tex : 1979) 2007;49:241–8.

37. Marín-García JJHFR. Mitochondrial DNA repair: a novel therapeutic target for heart failure. 2016;21:475–87.

38. Munzel T, Camici GG, Maack C, Bonetti NR, Fuster V, Kovacic JC. Impact of Oxidative Stress on the Heart and Vasculature: Part 2 of a 3-Part Series. Journal of the American College of Cardiology 2017;70:212–29.

39. Zorov DB, Juhaszova M, Sollott SJ. Mitochondrial reactive oxygen species (ROS) and ROS-induced ROS release. Physiological reviews 2014;94:909–50.

40. Cline SD. Mitochondrial DNA damage and its consequences for mitochondrial gene expression. Biochimica et biophysica acta 2012;1819:979–91.

41. Yakes FM, Van Houten B. Mitochondrial DNA damage is more extensive and persists longer than nuclear DNA damage in human cells following oxidative stress. Proceedings of the National Academy of Sciences of the United States of America 1997;94:514–9.

42. Casademont J, Miro O. Electron transport chain defects in heart failure. Heart failure reviews 2002;7:131–9.

43. Verdejo HE, del Campo A, Troncoso R, et al. Mitochondria, myocardial remodeling, and cardiovascular disease. Current hypertension reports 2012;14:532–9.

44. Madamanchi NR, Vendrov A, Runge MS. Oxidative stress and vascular disease. Arteriosclerosis, thrombosis, and vascular biology 2005;25:29–38.

45. Guzik TJ, Touyz RM. Oxidative Stress, Inflammation, and Vascular Aging in Hypertension. Hypertension (Dallas, Tex : 1979) 2017;70:660–7.

46. Lewis W. Defective mitochondrial DNA replication and NRTIs: pathophysiological implications in AIDS cardiomyopathy. American journal of physiology Heart and circulatory physiology 2003;284:H1–9.

47. Nitiss KC, Nitiss JL. Twisting and ironing: doxorubicin cardiotoxicity by mitochondrial DNA damage. Clinical cancer research : an official journal of the American Association for Cancer Research 2014;20:4737–9.

48. Lebrecht D, Walker UA. Role of mtDNA lesions in anthracycline cardiotoxicity. Cardiovascular toxicology 2007;7:108–13.

49. Knez J, Lakota K, Bozic N, et al. Correlation Between Mitochondrial DNA Content Measured in Myocardium and Peripheral Blood of Patients with Non-Ischemic Heart Failure. Genetic testing and molecular biomarkers 2017;21:736–41.

